# Evidence for a detection ceiling in the 2026 Bundibugyo virus disease epidemic in the Democratic Republic of the Congo: an analysis of publicly reported aggregate surveillance data

**DOI:** 10.64898/2026.07.16.26358218

**Authors:** Oussama Wael Bouhentala

## Abstract

**Background:** The 2026 Bundibugyo virus disease (BVD) epidemic in the Democratic Republic of the Congo (DRC) was declared on 15 May 2026 and determined a public health emergency of international concern on 17 May 2026. Public surveillance reporting consists of cumulative counts by report date; no line list with symptom-onset dates is available. Widely circulated characterisations — that this is the fastest-growing Ebola outbreak on record, that reported cases are doubling every 22 days, and that the case fatality ratio (CFR) is 37.5% — rest on these aggregates. We examined what the published data actually support.

**Methods:** We assembled twelve published anchor points (15 May–13 July 2026) from WHO, WHO AFRO, NICD and the DRC Institut National de Santé Publique; one was recovered by back-calculation and checked against the directly reported subsequent total. We computed mean daily incidence between anchors and the within-interval death-to-case ratio. We reconstructed symptom-onset dates by Richardson–Lucy deconvolution with right-truncation correction under assumed onset-to-report delays (mean 5, 7 and 9 days), estimated the instantaneous reproduction number (Cori method), and computed three CFR estimators: crude, resolved-case, and outcome-delay-adjusted. Provincial CFRs used exact binomial intervals.

**Results:** Confirmed cases plateaued at 40–52 per day for eighteen days (25 June–13 July). Over the same period the within-interval death-to-case ratio rose from 0.28 to 0.58. Reconstructed Rt was 1.28 (95% CrI 1.15–1.41) on 7 July, having fallen from approximately 2.9 in mid-May. Growth on the reconstructed onset curve corresponded to a doubling time of approximately 90 days, against 22 days computed from cumulative counts. CFR estimates were 37.5% (crude), 50.2% (outcome-delay-adjusted) and 67.3% (resolved-case). Crude provincial CFR was 34.9% (95% CI 32.7–37.1) in Ituri, 58.2% (50.7–65.5) in North Kivu and 81.0% (58.1–94.6) in newly affected provinces.

**Conclusions:** A flat case count accompanied by a rising death-to-case ratio is difficult to reconcile with a transmission plateau and is consistent with saturated case detection. Reported case counts appear to have substantially decoupled from transmission, and cannot presently distinguish control from detection failure. Doubling times computed from cumulative totals are artefacts. Test volume and positivity by health zone are the critical missing denominators.

## Introduction

Bundibugyo virus (BDBV, species Orthoebolavirus bundibugyoense) is one of four orthoebolaviruses known to cause disease in humans. It was first identified in Bundibugyo District, Uganda, in 2007 [1], and caused a second recognised outbreak in Isiro, Democratic Republic of the Congo (DRC), in 2012 [2]. Unlike Zaire ebolavirus, BDBV has no licensed vaccine and no approved specific therapeutic; the vaccine used in recent Zaire ebolavirus responses is not licensed against BDBV, and WHO has judged the available cross-protection evidence insufficient to support emergency ring vaccination in the current event [3].

The DRC declared the third recognised BDBV outbreak globally — and its seventeenth Ebola disease outbreak overall — on 15 May 2026, following laboratory confirmation and genomic classification by the Institut National de Recherche Biomédicale [4]. Uganda declared a linked outbreak the same day. The WHO Director-General determined the event a public health emergency of international concern on 17 May 2026 [5]. By 13 July 2026, the DRC had reported 2,011 laboratory-confirmed cases and 754 confirmed deaths across 45 health zones in five provinces [22].

Public surveillance reporting for this event consists almost entirely of cumulative counts by report date. No line list with symptom-onset dates has been released, and neither test volume nor test positivity has been published. Yet several quantitative characterisations have entered wide circulation and are shaping the interpretation of the epidemic: that it is the fastest-growing Ebola outbreak WHO has managed; that reported cases are doubling approximately every 22 days; and that the case fatality ratio is 37.5%.

Each of these claims is derived from the same cumulative aggregates, and each carries a specific analytical vulnerability. Doubling times computed from cumulative totals are not epidemic growth rates. Crude CFR computed during an epidemic in progress is downward-biased by unresolved cases. And any inference from reported case counts presumes that ascertainment is stable enough for those counts to track transmission — a presumption that is rarely tested and, in this setting, is questionable: as of 13 July, only 67.4% of identified case contacts were under follow-up across Ituri, North Kivu and Haut-Uele, while 92.3% of 430 investigated deaths through 5 July occurred in the community or before admission [21,22].

We therefore asked three questions of the published data. First, what is the epidemic’s actual growth rate, computed correctly? Second, what range of case fatality is defensible? Third — and most consequentially — do reported case counts still carry information about transmission, or has case ascertainment become the binding constraint on what is observed?

## Methods

### Data sources

We assembled twelve anchor points describing cumulative laboratory-confirmed cases, deaths and (where available) recoveries in the DRC between 15 May and 13 July 2026, from WHO Disease Outbreak News [4,6,7], WHO AFRO Weekly External Situation Reports [8–10,21,23], a NICD situational update [11], and the DRC Institut National de Santé Publique SitRep 060 [22]. Only DRC laboratory-confirmed cases were analysed. Anchors are listed in Table 1.

**Table 1.** Anchor series: cumulative laboratory-confirmed cases, deaths and recoveries, Democratic Republic of the Congo, 15 May–13 July 2026.

| Date (data as of) | Cases | Deaths | Recovered | Source |
| --- | --- | --- | --- | --- |
| 15 May | 8 | 4 | — | WHO PHEIC statement [5] |
| 21 May | 83 | 9 | — | WHO DON603 [4], excluding Uganda |
| 31 May | 321 | 48 | — | WHO AFRO SitRep 03 [23] |
| 7 Jun | 550 | 101 | — | WHO AFRO SitRep 04 [8] |
| 14 Jun | 808 | 192 | — | WHO AFRO SitRep 05 [9] |
| 17 Jun | 896 | 232 | — | WHO DON608 [6] |
| 25 Jun | 1,155 | 304 | 138 | NICD [11] |
| 28 Jun | 1,305 | 377 | — | WHO AFRO SitRep 08 [10], back-calculated |
| 1 Jul | 1,460 | 452 | — | WHO DON612 [7] |
| 5 Jul | 1,622 | 521 | — | WHO AFRO SitRep 08 [10] |
| 12 Jul | 1,963 | 719 | 333 | WHO AFRO SitRep 09 [21] |
| 13 Jul | 2,011 | 754 | 366 | INSP DRC SitRep 060 [22] |

One anchor was not published directly and was recovered by back-calculation from reported percentage changes. WHO AFRO Situation Report 08 reported increments of +317 cases (+24.3%) and +144 deaths (+38.2%) since Situation Report 07, giving 317/0.243 = 1,305 cases and 144/0.382 = 377 deaths at 28 June after rounding to whole counts. Adding the reported increments gives 1,622 cases and 521 deaths on 5 July, exactly matching the totals reported directly in Situation Report 08. The independently published crude CFRs of 28.8% and 32.1% are also reproduced to within 0.1 percentage points, supporting internal consistency.

### Reported incidence between anchors

To obtain a minimally modelled description of epidemic speed, we computed the mean daily increment in confirmed cases and deaths between consecutive anchors, and the ratio of deaths to cases accrued within each interval. The only assumption is that counts are distributed evenly between anchors. Intervals are unequal (1–10 days) and short intervals are correspondingly noisy.

### Reconstruction of symptom-onset dates

For modelled analyses, the cumulative curve was interpolated onto a daily grid using monotone piecewise cubic Hermite interpolation and differenced to give daily reported incidence. This preserves monotonicity and reproduces every anchor exactly, but invents between-anchor shape.

Onset dates were reconstructed by Richardson–Lucy expectation-maximisation deconvolution with right-truncation correction, following Goldstein et al. [12]. The truncation normaliser *q*_s_ is the probability that a case with onset on day *s* has been reported by the data cut-off; division by it inflates recent onsets and constitutes the nowcast. Iteration was stopped at 30 (χ^2^/df ≈ 0.03; the reconstructed tail was stable across 20–40 iterations). Onset estimates are presented as reliable where *q*_s_ ≥ 0.50, provisional where 0.20 ≤ *q*_s_ < 0.50, and are not shown below 0.20.

The onset-to-report delay was modelled as a gamma distribution with central assumption mean 7 days (SD 4.5), with sensitivity scenarios at mean 5 and 9 days. No BDBV-specific onset-to-report distribution has been published; these are informed assumptions and constitute the largest single source of uncertainty in the modelled analyses. Uncertainty intervals pool 300 Poisson resamples across the three delay scenarios and are reported as 2.5th–97.5th percentiles.

### Reproduction number

The instantaneous reproduction number was estimated from the reconstructed onset curve using the method of Cori et al. [13], with a 7-day sliding window and a Gamma(1, 5) prior. The serial interval was taken as gamma with mean 15.3 days and SD 9.3 days, borrowed from Zaire ebolavirus [14]; no BDBV-specific estimate has been published. Estimation from onset dates approximates, but does not equal, estimation from infection dates; no incubation-period deconvolution was applied.

### Case fatality estimators

Three estimators were computed. The crude ratio is deaths divided by all confirmed cases, *D/C*. The resolved-case ratio is *D/(D+R)*. The outcome-delay-adjusted ratio, following Nishiura et al. [15] and Ghani et al. [16], is *D/(C·ut)*, where *ut* is the proportion of reported cases expected to have a known outcome by day *t*. Report-to-outcome delays were modelled as gamma with mean 6 days (SD 4) for death and mean 16 days (SD 6) for recovery; the outcome mix was solved by fixed-point iteration. Provincial CFRs are crude, with exact binomial (Clopper–Pearson) intervals [17]. Province-specific counts for South Kivu, Haut-Uele and Tshopo were verified directly against the 13 July national situation report and pooled for the combined stratum [22].

### Software and reproducibility

Analyses used Python 3 with NumPy, SciPy, pandas and Matplotlib. The full pipeline is deterministic (seed 20260715), The analysis code, anchor dataset and derived series will be deposited in a public repository before journal submission.

### Use of artificial intelligence

In accordance with the ICMJE Recommendations (January 2026 revision), the following is disclosed. A large language model (Claude, Anthropic) was used interactively to assist with source retrieval, to write the analysis code implementing the deconvolution, reproduction-number and case-fatality estimators, to generate the figures from that code, and to draft and edit manuscript text. The choice of methods, the interpretation, and all claims made in this paper are the author’s, who has verified the analysis, independently checked the outputs, and takes full responsibility for the content. No AI system is an author. The figures are programmatic renderings of the analysed data; none is a generative image. Readers and reviewers are directed to the deposited code, which permits full independent verification of every computational step.

### Ethics

This analysis used only publicly available, aggregate surveillance data containing no personal or identifying information. It did not involve human participants and required no ethical approval or informed consent.

## Results

### Reported incidence has plateaued

The anchor series is given in Table 1. Mean daily confirmed cases rose from 12.5 per day in the interval ending 21 May to approximately 33 per day through June, then to a plateau of 40–52 per day sustained across the five reporting intervals ending 28 June, 1 July, 5 July, 12 July and 13 July — eighteen days in total (Figure 1A). Over the same period, mean daily confirmed deaths continued to rise, from 9.0 per day in the interval ending 25 June to 28.3 per day in the interval ending 12 July.

**Figure 1.**
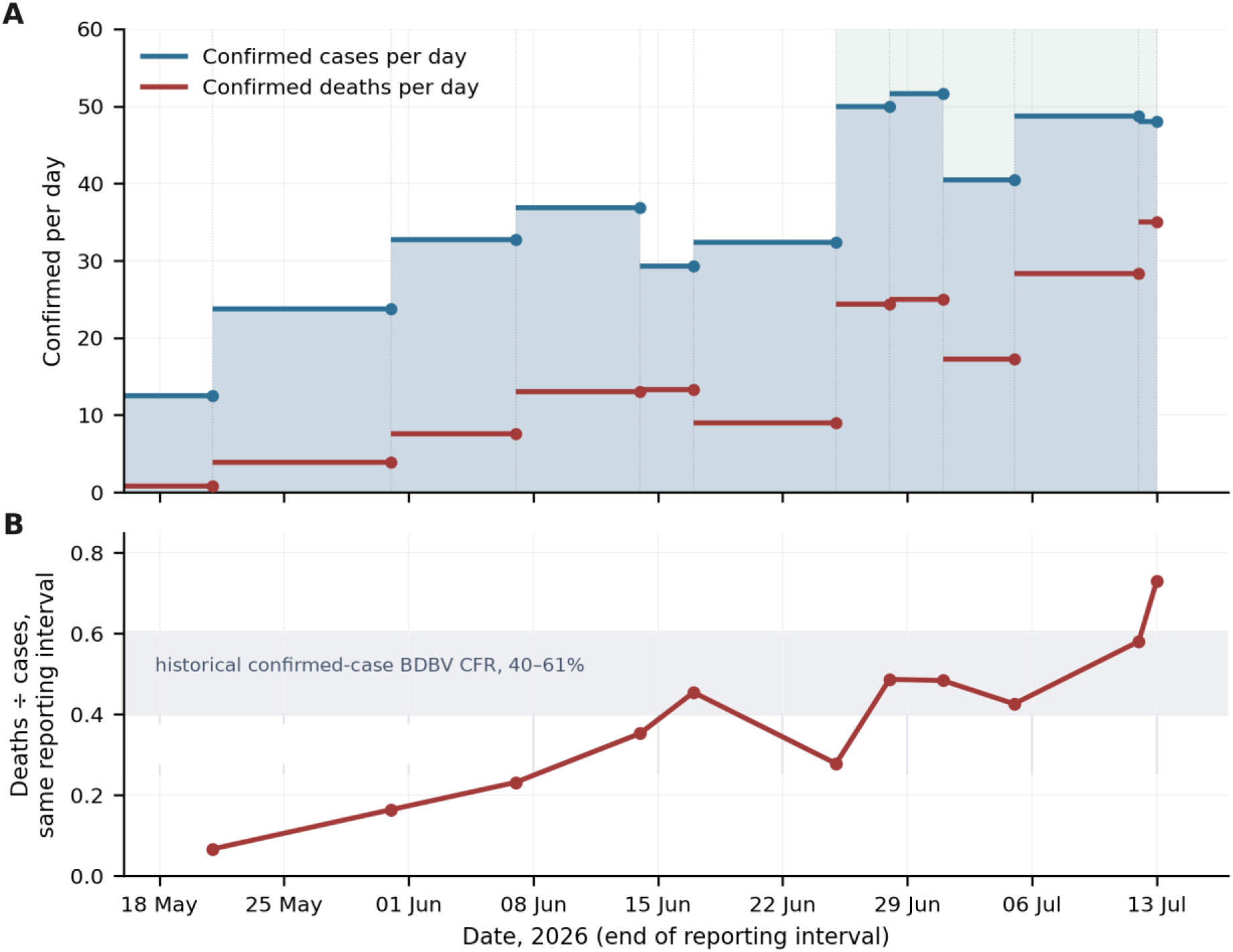
Reported incidence and the death-to-case ratio, DRC confirmed cases, 15 May–13 July 2026. (A) Mean daily confirmed cases (blue) and deaths (red) between consecutive published anchors; shading marks the 40–52 cases/day plateau from 25 June. (B) Ratio of deaths to cases accrued within the same reporting interval; grey band shows the historical laboratory-confirmed BDBV CFR range (approximately 40–61%). Intervals are unequal (1–10 days); the final point rests on a single day and is correspondingly noisy. Minimal modelling: the only assumption is even distribution of counts between anchors.

### The death-to-case ratio is rising against a flat case count

The ratio of deaths to cases accrued within each reporting interval rose monotonically across four consecutive intervals, from 0.28 (ending 25 June) through 0.49, 0.48 and 0.43 to 0.58 (ending 12 July), reaching 0.73 in the single-day interval ending 13 July (Figure 1B). By mid-July, each reporting interval was yielding roughly three deaths for every five cases detected.

### Reconstructed onset curve and reproduction number

Deconvolution under the central delay assumption placed the reconstructed peak of symptom onset around 24 June at approximately 77 cases per day, declining to approximately 56 per day by 7 July, the last date at which at least half of onsets would be expected to have been reported (Figure 2A). An estimated 331 cases (95% CI 100–554) had experienced onset but had not yet been reported at the data cut-off.

**Figure 2.**
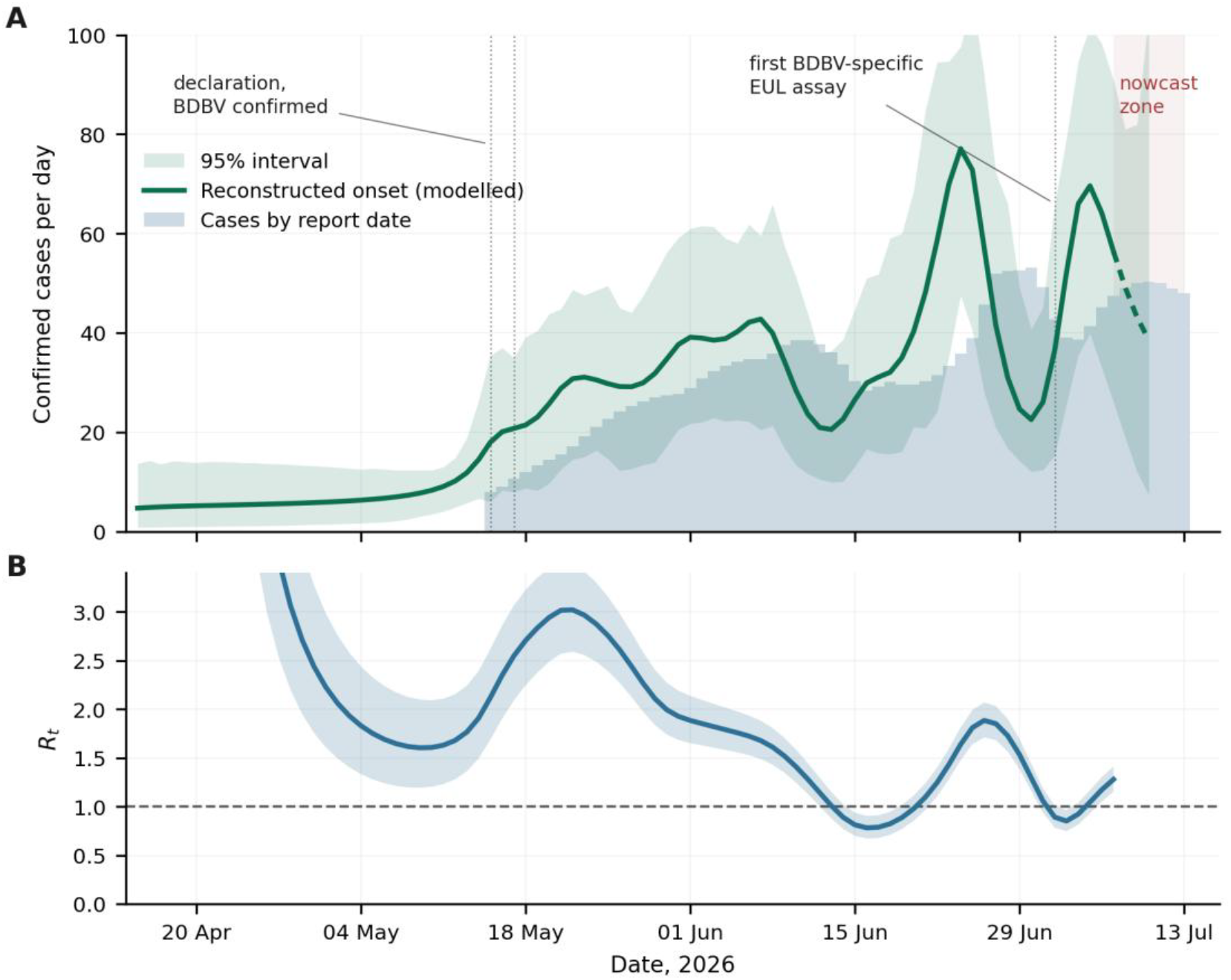
Modelled reconstruction of symptom-onset timing and transmission. (A) Cases by reconstructed date of symptom onset (green line; dashed where fewer than 50% of onsets are expected to have been reported) with 95% interval pooling three delay scenarios and 300 Poisson resamples, against cases by report date (bars). (B) Instantaneous reproduction number with 95% credible interval. The left tail does not represent the true early epidemic: deconvolution can only shift back cases that were eventually confirmed, and transmission before 15 May was never laboratory-confirmed.

The instantaneous reproduction number fell from 2.94 (95% CrI 2.49–3.45) on 20 May to 1.89 (1.65– 2.14) on 1 June, 1.41 (1.25–1.58) on 10 June, and thereafter plateaued slightly above unity: 1.30 (1.16–1.44) on 30 June and 1.28 (1.15–1.41) on 7 July (Figure 2B). This trajectory was robust across the three delay scenarios. The excursion below 1 around 20 June and the subsequent rebound track anchor spacing and should not be interpreted.

Growth on the reconstructed onset curve over the final fourteen reliable days corresponded to r = 0.008 per day, a doubling time of approximately 90 days. The equivalent figure computed from cumulative confirmed cases over 17 June–13 July (896 to 2,011) is 22 days — a fourfold discrepancy arising because a cumulative total doubles even when incidence is constant.

### Case fatality is an interval, not a point

At the data cut-off, 74.7% of reported cases would be expected to have a known outcome. The three estimators gave 37.5% (crude), 50.2% (outcome-delay-adjusted) and 67.3% (resolved-case) (Table 2, Figure 3A). The crude and resolved-case values bound the estimate from below and above respectively; the adjusted value is the most plausible point estimate.

**Table 2.** Case fatality ratio among laboratory-confirmed cases, DRC, 13 July 2026, by estimator.

| Estimator | Estimate | Property |
| --- | --- | --- |
| Crude, D/C | 37.5% | Lower bound; denominator contains cases of unknown outcome |
| Outcome-delay-adjusted, $D/(C \cdot ut)$ | 50.2% | Divides by $ut = 74.7\%$, the proportion of cases expected to have a known outcome; most plausible point estimate |
| Resolved-case, $D/(D+R)$ | 67.3% | Upper bound; deaths resolve faster (mean 6 d) than recoveries (mean 16 d) |

**Figure 3.**
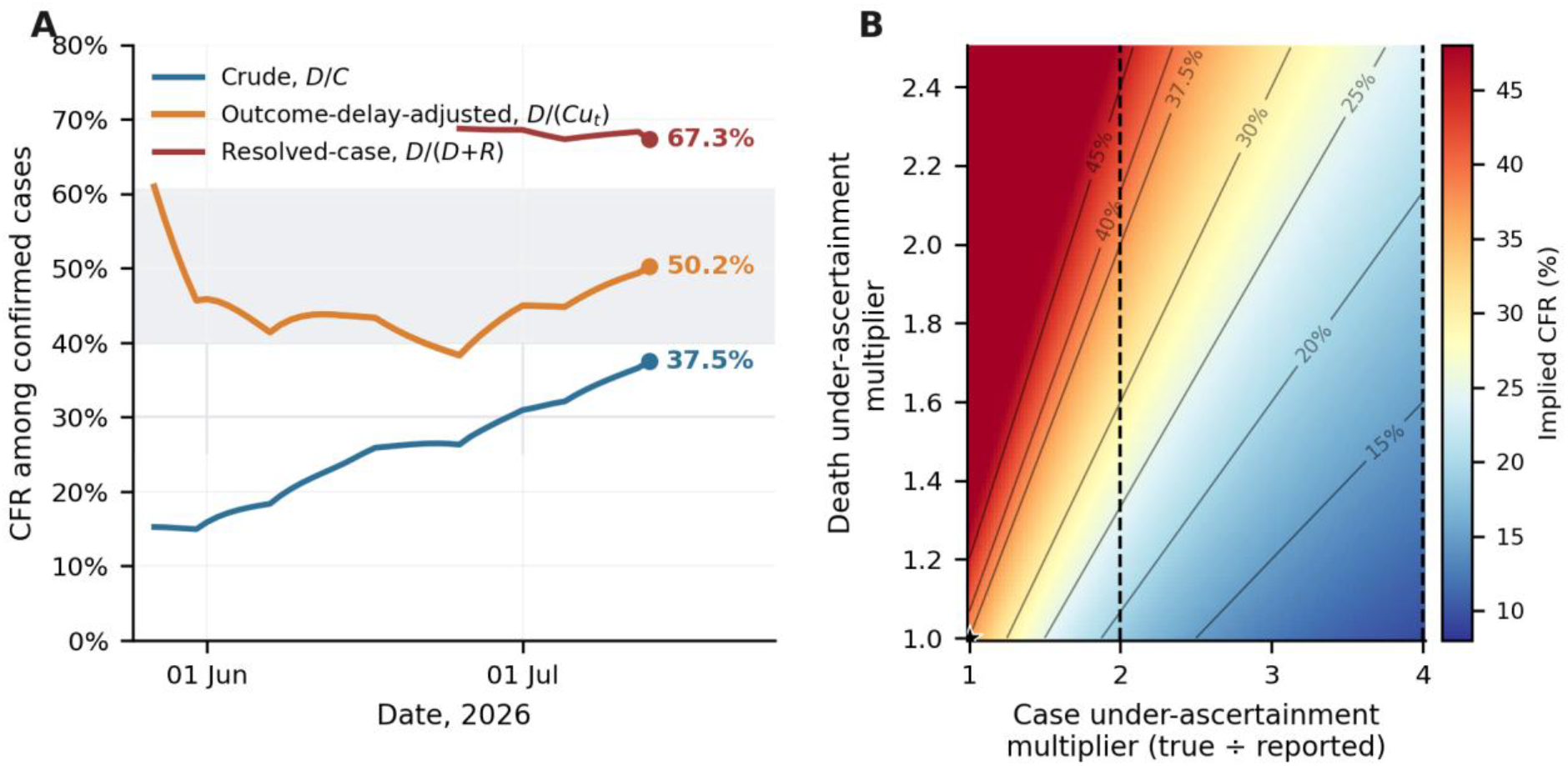
Case fatality among laboratory-confirmed cases. (A) Three estimators over time; grey band shows the historical laboratory-confirmed BDBV CFR range (approximately 40–61%). (B) Implied CFR as a function of case under-ascertainment (horizontal) and death under-ascertainment (vertical), holding the 13 July reported values fixed (754 deaths, 2,011 cases); star marks complete ascertainment of both; dashed lines illustrate two- and fourfold case under-ascertainment scenarios. The two biases act in opposite directions.

Historical BDBV CFR estimates depend strongly on case definition and outcome availability. Towner et al. reported 37 deaths among 149 suspected cases (24.8%) and also described a preliminary estimate of approximately 36%; MacNeil et al. observed 17 deaths among 43 laboratory-confirmed patients with an acute-phase sample and known outcome (39.5%); and Kratz et al. reported 22 deaths among 36 confirmed cases (61.1%) [1,2,19]. For comparison with the present laboratory-confirmed series, the most defensible historical confirmed-case range is therefore approximately 40–61%, within which the adjusted estimate of 50.2% lies. Figure 3B shows that case and death under-ascertainment move the implied CFR in opposite directions; without independent ascertainment data, CFR alone cannot identify the magnitude of missed infections.

### A steep provincial gradient in crude case fatality

Crude CFR was 34.9% (95% CI 32.7–37.1) in Ituri, which accounted for 89.9% of confirmed cases; 58.2% (50.7–65.5) in North Kivu; and 81.0% (58.1–94.6) in South Kivu, Haut-Uele and Tshopo combined (Table 3, Figure 4). The intervals for Ituri and North Kivu do not overlap. The third stratum rests on 21 cases and its interval is correspondingly wide.

**Table 3.**
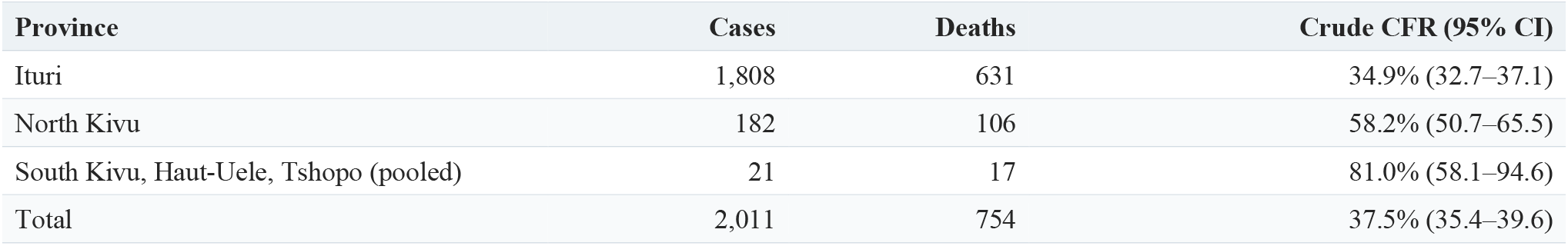
Provincial burden and crude case fatality with exact binomial confidence intervals, DRC confi[-rmed cases, 13 July 2026. The pooled third stratum comprises South Kivu (3 cases, 1 death), Haut-Uele (14 cases, 13 deaths) and Tshopo (4 cases, 3 deaths).

| Province | Cases | Deaths | Crude CFR (95% CI) |
| --- | --- | --- | --- |
| Ituri | 1,808 | 631 | 34.9% (32.7–37.1) |
| North Kivu | 182 | 106 | 58.2% (50.7–65.5) |
| South Kivu, Haut-Uele, Tshopo (pooled) | 21 | 17 | 81.0% (58.1–94.6) |
| Total | 2,011 | 754 | 37.5% (35.4–39.6) |

**Figure 4.**
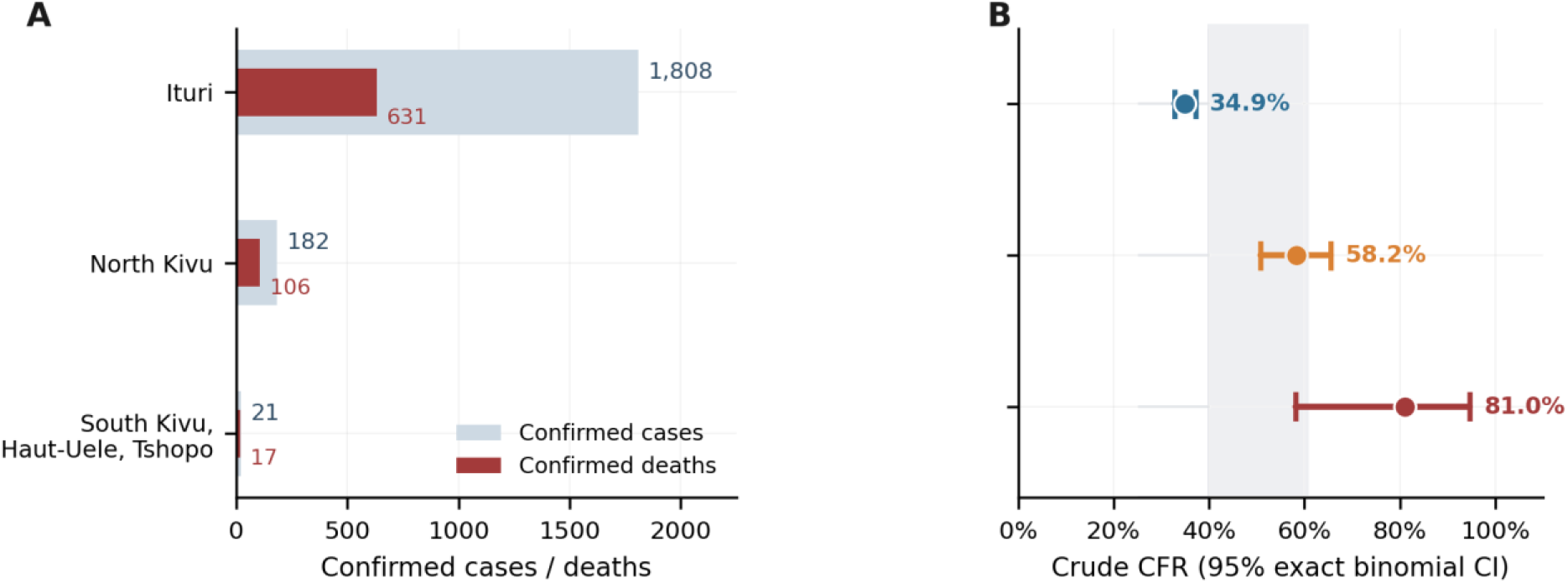
Provincial burden and crude case fatality, DRC confirmed cases, 13 July 2026. (A) Confirmed cases and deaths by province. (B) Crude CFR with 95% exact binomial intervals; grey band shows the historical laboratory-confirmed BDBV CFR range (approximately 40–61%). Crude CFR is a lower bound in every stratum. The pooled third stratum comprises South Kivu (3 cases, 1 death), Haut-Uele (14 cases, 13 deaths) and Tshopo (4 cases, 3 deaths), verified directly against the national situation report [22].

## Discussion

Reported confirmed cases in the DRC have been flat for eighteen days while the proportion of detected cases that are fatal has risen steadily. These two observations are difficult to reconcile with a genuine plateau in transmission. Under a transmission plateau, deaths would be expected to flatten one to two weeks after cases, reflecting the interval from detection to outcome, and the within-interval death-to-case ratio would stabilise. Instead it climbed across four consecutive intervals.

The pattern is what would be expected if case detection had saturated: a system finding a roughly fixed number of cases per day, and finding them increasingly at the severe end of the clinical spectrum. This interpretation is consistent with the independent observations that, as of 13 July, 67.4% of identified case contacts were under follow-up across the three provinces reporting active contact monitoring, and that 92.3% of 430 investigated deaths through 5 July occurred in the community or before admission [21,22]. It is reinforced by the provincial gradient: crude CFR rises steeply from the epicentre outward, and the same virus is circulating throughout, so a 23-point difference in apparent lethality across a provincial boundary is more plausibly a gradient in detection and treatment access than in biology.

The central implication is uncomfortable. A plateau produced by saturated detection is indistinguishable, in a cumulative case count, from a plateau produced by successful control. If this interpretation is correct, the response’s headline indicator has stopped discriminating between success and failure, and a subsequent fall in reported cases would be as consistent with deteriorating ascertainment as with epidemic decline.

### Two claims that the data do not support

First, doubling times computed from cumulative counts are artefacts. The widely quoted 22 days differs from the correctly computed value by a factor of four, and the two point to opposite operational conclusions. This is an elementary error, but it has evidently propagated, and it should be retired from discussion of this epidemic.

Second, the reconstruction does not support the characterisation of this as an unusually fast epidemic. An Rt near 1.3 represents sustained growth but sits well below the 1.7–2.0 estimated during the explosive phase of the 2014 West Africa epidemic [14]. The distinction matters operationally: an epidemic that is fast requires surge capacity, whereas an epidemic that is invisible requires case-finding. The language of explosive growth may have directed attention toward the former when the evidence points to the latter.

### CFR does not identify the ascertainment mechanism

The historical comparison does not provide an independent estimate of the case undercount. The current outcome-delay-adjusted estimate lies within the approximately 40–61% range reported among laboratory-confirmed cases in the two previous outbreaks [2,19]. Figure 3B instead demonstrates non-identifiability: case under-ascertainment lowers the implied CFR, whereas death under-ascertainment raises it. The elevated crude CFR in some provinces remains compatible with late detection, restricted treatment access, outcome maturation and differential confirmation of deaths, but it is not by itself a quantitative estimate of missed infections.

Uganda provides an informative natural comparison. Facing the same virus, with the same absence of a licensed vaccine and approved therapeutic, Uganda reported 20 confirmed cases — 15 imported and five secondary — with no sustained tertiary transmission and no confirmed case after 21 June [7]. Uganda’s transmission network remained visible; the DRC’s largely does not. This is the strongest available indication that the DRC epidemic is not biologically intractable.

### Limitations

This analysis is constrained by what has been published, and its central argument — that the published data have stopped being informative about transmission — applies to the analysis itself. We had no access to line lists, laboratory registers, contact-tracing databases or field observation.

The interpolation between anchors invents daily structure. Real reporting is lumpy, and WHO has warned that batch confirmation of backlogged samples inflates single-day counts; the method cannot distinguish backlog from incident transmission. Some of the late-June rise in reported incidence is likely attributable to laboratory expansion and to the first BDBV-specific assay, which received WHO Emergency Use Listing on 2 July [18] — meaning ascertainment was not constant across the series, a violation the uncertainty intervals do not capture. This cuts both ways: improving ascertainment during the plateau would make the underlying transmission plateau more real than it appears, whereas deteriorating ascertainment would make it less so. The rising death-to-case ratio favours the latter, but does not establish it.

The delay distributions are assumed rather than estimated, and the serial interval is borrowed from a different orthoebolavirus; if the BDBV serial interval is shorter, Rt is overestimated. The left tail of the reconstruction cannot represent pre-declaration transmission, which was not included in the laboratory-confirmed anchor series and is structurally unrecoverable. The final death-to-case ratio rests on a single day. The pooled third provincial stratum rests on only 21 cases, although its component province counts were verified directly against the national situation report [22]. Population denominators were not applied, because reliable current denominators are unavailable amid mass displacement, so no attack rates are reported. Finally, an alternative and non-exclusive explanation for both the provincial gradient and the rising death-to-case ratio is deteriorating access to supportive care in conflict-affected areas; this would not be distinguishable from a pure detection effect using these data, though both point to the same operational conclusion.

### Implications

The single most valuable missing datum is test volume and test positivity by health zone over time. Without a denominator, a case plateau cannot be interpreted. Flat positivity on flat testing would indicate a capacity ceiling; rising positivity on flat testing would indicate transmission outrunning a fixed detection capacity, which the death-to-case trajectory suggests. These data already exist in laboratory registers and their publication would be a low-cost, high-value change.

For monitoring, the proportion of new cases already registered as contacts before symptom onset is the indicator that should be prioritised, because — unlike the case count — it is not a function of testing volume and can therefore distinguish a detection ceiling from genuine control. A sustained transition from most cases arising from unknown chains to most cases arising among monitored contacts would be the first credible evidence that the epidemic is turning.

## Conclusion

Reported case counts in the 2026 DRC Bundibugyo epidemic appear to have substantially decoupled from transmission. The epidemic is not, on this evidence, unusually fast; it is unusually invisible. Recognising the difference matters, because it determines whether the response invests in capacity or in case-finding, and because it determines whether the numbers currently being watched can tell anyone whether either is working.

## Data Availability

All data analysed in the present study were obtained from publicly available aggregate surveillance reports published by the World Health Organization, WHO Regional Office for Africa, the European Centre for Disease Prevention and Control, the National Institute for Communicable Diseases, and the Democratic Republic of the Congo health authorities. The source reports are cited in the manuscript. The extracted data used for the analysis are presented in Table 1, and all source reports are cited in the manuscript.

## Declarations

### Ethics approval

Not required; the analysis used only publicly available aggregate data containing no personal information.

### Data and code availability

**The analysis code, anchor dataset and derived series will be deposited in a public repository before journal submission**.. **All source data are publicly available from the cited WHO, WHO AFRO, INSP DRC and NICD reports**.

### Use of AI

Disclosed in the Methods, in accordance with the ICMJE Recommendations (January 2026).

### Author contributions

OWB conceived the analysis, specified the methods, verified the outputs, interpreted the results, drafted and revised the manuscript, and takes responsibility for its content.

## Acknowledgements

The author thanks the responders of the DRC and Ugandan Ebola responses, whose reporting under conditions of armed conflict made this analysis possible.

## References

1. Towner JS, Sealy TK, Khristova ML, Albariño CG, Conlan S, Reeder SA, et al. Newly discovered Ebola virus associated with hemorrhagic fever outbreak in Uganda. PLoS Pathog. 2008;4(11):e1000212. doi:10.1371/journal.ppat.1000212.

2. Kratz T, Roddy P, Tshomba Oloma A, Jeffs B, Pou Ciruelo D, de la Rosa O, et al. Ebola virus disease outbreak in Isiro, Democratic Republic of the Congo, 2012: signs and symptoms, management and outcomes. PLoS One. 2015;10(6):e0129333. doi:10.1371/journal.pone.0129333.

3. World Health Organization. Ebola vaccines. Questions and answers. 25 June 2026. Available from: https://www.who.int/news-room/questions-and-answers/item/ebola-vaccines

4. World Health Organization. Ebola disease caused by Bundibugyo virus – Democratic Republic of the Congo. Disease Outbreak News, DON603. 21 May 2026. Available from: https://www.who.int/emergencies/disease-outbreak-news/item/2026-DON603

5. World Health Organization. Epidemic of Ebola disease caused by Bundibugyo virus in the Democratic Republic of the Congo and Uganda determined a public health emergency of international concern. 17 May 2026. Available from: https://www.who.int/news/item/17-05-2026-epidemic-of-ebola-disease-in-the-democratic-republic-of-the-congo-and-uganda-determined-a-public-health-emergency-of-international-concern

6. World Health Organization. Bundibugyo virus disease, Democratic Republic of the Congo and Uganda. Disease Outbreak News, DON608. 19 June 2026. Available from: https://www.who.int/emergencies/disease-outbreak-news/item/2026-DON608

7. World Health Organization. Ebola disease caused by Bundibugyo virus, Democratic Republic of the Congo and Uganda. Disease Outbreak News, DON612. 3 July 2026. Available from: https://www.who.int/emergencies/disease-outbreak-news/item/2026-DON612

8. World Health Organization Regional Office for Africa. Ebola Bundibugyo virus disease outbreak, Democratic Republic of the Congo and Uganda: Weekly External Situation Report 04, data as of 7 June 2026. Available from: https://www.afro.who.int/countries/uganda/publication/ebola-bundibugyo-virus-disease-outbreak-democratic-republic-congo-0

9. World Health Organization Regional Office for Africa. Ebola Bundibugyo virus disease outbreak, Democratic Republic of the Congo and Uganda: Weekly External Situation Report 05, data as of 14 June 2026. Available from: https://www.afro.who.int/countries/uganda/publication/ebola-bundibugyo-virus-disease-outbreak-democratic-republic-congo-1

10. World Health Organization Regional Office for Africa. Ebola Bundibugyo virus disease outbreak, Democratic Republic of the Congo and Uganda: Weekly External Situation Report 08, data as of 5 July 2026. Available from: https://www.afro.who.int/countries/democratic-republic-of-congo/publication/ebola-bundibugyo-virus-disease-outbreak-1

11. National Institute for Communicable Diseases. Situational update on the Ebola disease outbreak caused by Bundibugyo virus (26 June 2026). Published 29 June 2026. Available from: https://www.nicd.ac.za/situational-update-on-the-ebola-disease-outbreak-caused-by-bundibugyo-virus/

12. Goldstein E, Dushoff J, Ma J, Plotkin JB, Earn DJD, Lipsitch M. Reconstructing influenza incidence by deconvolution of daily mortality time series. Proc Natl Acad Sci U S A. 2009;106(51):21825–21829. doi:10.1073/pnas.0902958106.

13. Cori A, Ferguson NM, Fraser C, Cauchemez S. A new framework and software to estimate time-varying reproduction numbers during epidemics. Am J Epidemiol. 2013;178(9):1505–1512. doi:10.1093/aje/kwt133.

14. WHO Ebola Response Team. Ebola virus disease in West Africa — the first 9 months of the epidemic and forward projections. N Engl J Med. 2014;371(16):1481–1495. doi:10.1056/NEJMoa1411100.

15. Nishiura H, Klinkenberg D, Roberts M, Heesterbeek JAP. Early epidemiological assessment of the virulence of emerging infectious diseases: a case study of an influenza pandemic. PLoS One. 2009;4(8):e6852. doi:10.1371/journal.pone.0006852.

16. Ghani AC, Donnelly CA, Cox DR, Griffin JT, Fraser C, Lam TH, et al. Methods for estimating the case fatality ratio for a novel, emerging infectious disease. Am J Epidemiol. 2005;162(5):479–486. doi:10.1093/aje/kwi230.

17. Clopper CJ, Pearson ES. The use of confidence or fiducial limits illustrated in the case of the binomial. Biometrika. 1934;26(4):404–413. doi:10.1093/biomet/26.4.404.

18. World Health Organization. WHO adds first diagnostic test for Ebola Bundibugyo virus to its Emergency Use Listing. 2 July 2026. Available from: https://www.who.int/news/item/02-07-2026-who-adds-first-diagnostic-test-for-ebola-bundibugyo-virus-to-its-emergency-use-listing

19. MacNeil A, Farnon EC, Wamala J, Okware S, Cannon DL, Reed Z, et al. Proportion of deaths and clinical features in Bundibugyo Ebola virus infection, Uganda. Emerg Infect Dis. 2010;16(12):1969–1972. doi:10.3201/eid1612.100627.

20. International Committee of Medical Journal Editors. Recommendations for the conduct, reporting, editing, and publication of scholarly work in medical journals. Updated January 2026. Available from: https://www.icmje.org/recommendations/

21. World Health Organization Regional Office for Africa. Ebola Bundibugyo virus disease outbreak, Democratic Republic of the Congo and Uganda: Weekly External Situation Report 09, data as of 12 July 2026. Available from: https://www.afro.who.int/countries/democratic-republic-of-congo/publication/ebola-bundibugyo-virus-disease-outbreak-2

22. Institut National de Santé Publique République démocratique du Congo. SitRep n°060: maladie à virus Bundibugyo, situation au 13 juillet 2026. Available from: https://insp.cd/sitrep-n060-mvb-13-07-2026/

23. World Health Organization Regional Office for Africa. Ebola Bundibugyo virus disease outbreak, Democratic Republic of the Congo and Uganda: Weekly External Situation Report 03, data as of 31 May 2026. Available from: https://www.afro.who.int/countries/uganda/publication/ebola-bundibugyo-virus-disease-outbreak-democratic-republic-congo-uganda-weekly-external-situation

